# Treatment of Six COVID-19 Patients with Convalescent Plasma

**DOI:** 10.1101/2020.05.21.20109512

**Authors:** Can Jin, Juan Gu, Youshu Yuan, Qinying Long, Qi Zhang, Hourong Zhou, Weidong Wu, Wei Zhang

## Abstract

**Objective:** To describe the efficacy of convalescent plasma transfusion for COVID-19 patients.

**Methods:** This is a retrospective study of 6 COVID-19 patients with convalescent plasma at Guizhou Provincial Jiangjunshan Hospital – a tertial hospital, in Guiyang, Guizhou, China, from January 29, to April 30, 2020; final data of follow-up was May 12, 2020. Through the review of the electronic medical records of Guizhou Jiangjunshan Hospital, clinical data of 6 patients were obtained. Three patients with worsening symptoms after empirical treatment with antivirals were transfused convalescent plasma therapy for the first treatment, while the other three severe or critical COVID-19 patients with rapid progression were transfused. The efficacy of convalescent plasma depends on the relief of symptoms, changes in laboratory indicators and chest imaging abnormalities.

**Results:** The PaO2 / FiO2 and lymphocyte count of patients 1, 2 and 3 treated with convalescent plasma treatment for the first treatment period were changed from abnormal to normal. The levels of inflammation markers CRP and IL-6 of the patients decreased significantly. Chest imaging examination showed that the lung lesions gradually subsided. The relapsed patients (No. 4 and No. 6), after using convalescent plasma therapy, turned negative on two consecutive throat swab tests on Day 24 and Day 3, respectively.

**Conclusions:** Convalescent plasma treatment of COVID-19 is beneficial for those patients with be difficult to turn to negative or re-positive RT-PCR.

**Key Points:** Convalescent plasma treatment of COVID-19 is beneficial for those patients with be difficult to turn to negative or re-positive RT-PCR.

## Introduction

In December 2019, Wuhan, China, became the center of pneumonia outbreaks caused by severe acute respiratory syndrome coronavirus 2 (SARS-CoV-2)[1]. The World Health Organization (WHO) subsequently named the condition coronavirus disease 2019 (COVID-19) and declared the outbreak a pandemic. As of June 6, 2020, the World Health Organization reported that globally, the number of COVID-19 confirmed cases reached 6,807,555, and the cumulative number of deaths reached 396,856. To date, no specific therapeutic agents or vaccines have been proven effective for COVID-19; supportive care and generic antiviral drugs are the only available treatments. Although several antiviral agents appear clinically beneficial, their efficacy is far from satisfactory, and their mechanisms of action remain unclear [2-4]. Ongoing research and clinical trials are investigating the effectiveness against COVID-19 of various experimental therapies, including Favipiravir, Lopinavir-ritonavir, Remdesivir, Chloroquine/Hydroxychloroquine, IL-6 inhibitors, and convalescent plasma [5].

Convalescent plasma is plasma obtained from a recovered individual after the infection has subsided and specific antibody has developed. Transfusion with convalescent plasma may reduce the clinical infection or the clinical severity in patients who have recently been infected with the same pathogen as the donor of the convalescent plasma [4]. Convalescent plasma therapy has a favorable effect on viral load, serum cytokine response, and mortality. Historically, there have been many successful applications of convalescent plasma therapy used as post-exposure prophylaxis (e.g., rabies, hepatitis, measles, mumps, and polio) and as treatment for various infectious diseases administered after infection (e.g., Ebola virus, Middle East respiratory syndrome, SARS-CoV, Argentine hemorrhagic fever, H5N1 avian influenza, and H1N1 influenza)[4-11]. Shen et al. recently reported the preliminary results of five critically ill patients with COVID-19 and acute respiratory distress syndrome (ARDS) who were treated with plasma from recovered individuals; they found that about one week after plasma transfusion, the clinical status of all patients improved, their body temperature returned to normal, and their Sequential Organ Failure Assessment scores and ratios of the partial pressure of oxygen (PaO_2_) to the fraction of inspired oxygen (FiO_2_) (PaO_2_ measured in mm Hg and FiO_2_ measured as the fraction of inspired oxygen) had improved as well. Within 12 days after the transfusion, the patient neutralizing antibody titers had increased, and their respiratory samples tested negative for SARS-CoV-2 [12].

These limited findings provide encouraging evidence that infusion with convalescent plasma may be beneficial for patients infected with SARS-CoV-2 [13]. As the current number of COVID-19 cases and the number of deaths related to the disease are increasing at an alarming rate, additional research to determine the safety and effectiveness of convalescent plasma therapy are urgently needed. The present work conducted a retrospective analysis to further assess the utility of convalescent plasma as a treatment for COVID-19.

## Methods

### Study design

This retrospective, single-center study was conducted on six COVID-19 patients who were treated with convalescent plasma at Guizhou Provincial Jiangjunshan Hospital in Guiyang, Guizhou, China from January 29, 2020 to April 30, 2020. The final follow-up data were collected on May 12, 2020. We obtained written informed consent from each participant. This study was approved by the Biomedical Ethics Committee of Affiliated Hospital of Zunyi Medical University and was registered with the Chinese Clinical Trial Registry Center. (CCTR number: ChiCTR 2000033056, registered 19 May 2020. URL: http://www.chictr.org.cn/edit.aspx?pid=53859&htm=4.)

### Patients

This study collected data from six patients admitted to Guizhou Jiangjunshan Hospital with laboratory confirmed COVID-19 cases based on positive SARS-CoV-2 real-time PCR results from throat swab samples who had undergone convalescent plasma transfusion. Our inclusion criteria were: patients with a SARS-CoV-2-positive laryngeal swab AND the rapid development of severe disease while SARS-CoV-2 RT-PCR throat swab results remained positive OR worsening symptoms after empirical treatment with antiviral drugs in recurrent disease (newly positive SARS-CoV-2 RT-PCR throat swab results after negative results had been obtained). The patients continued to use antiviral drugs while receiving convalescent plasma therapy.

### Clinical information

The clinical data of the six enrolled patients were acquired from a review of the hospital electronic medical records and included the following information: epidemiology, latency, symptoms, and treatment data. The baseline characteristics of the enrolled patients are shown in Table 1. The clinical indicators assessed before and after convalescent plasma transfusion included clinical data (PaO_2_/FiO_2_) and laboratory data (e.g., lymphocyte count, C-reactive protein (CRP) level, and IL-6 level), as well as any changes in disease complications. We also examined the length of time between the first SARS-CoV-2-positive laryngeal swab and the first following SARS-CoV-2-negative laryngeal swab.

**Table 1.**
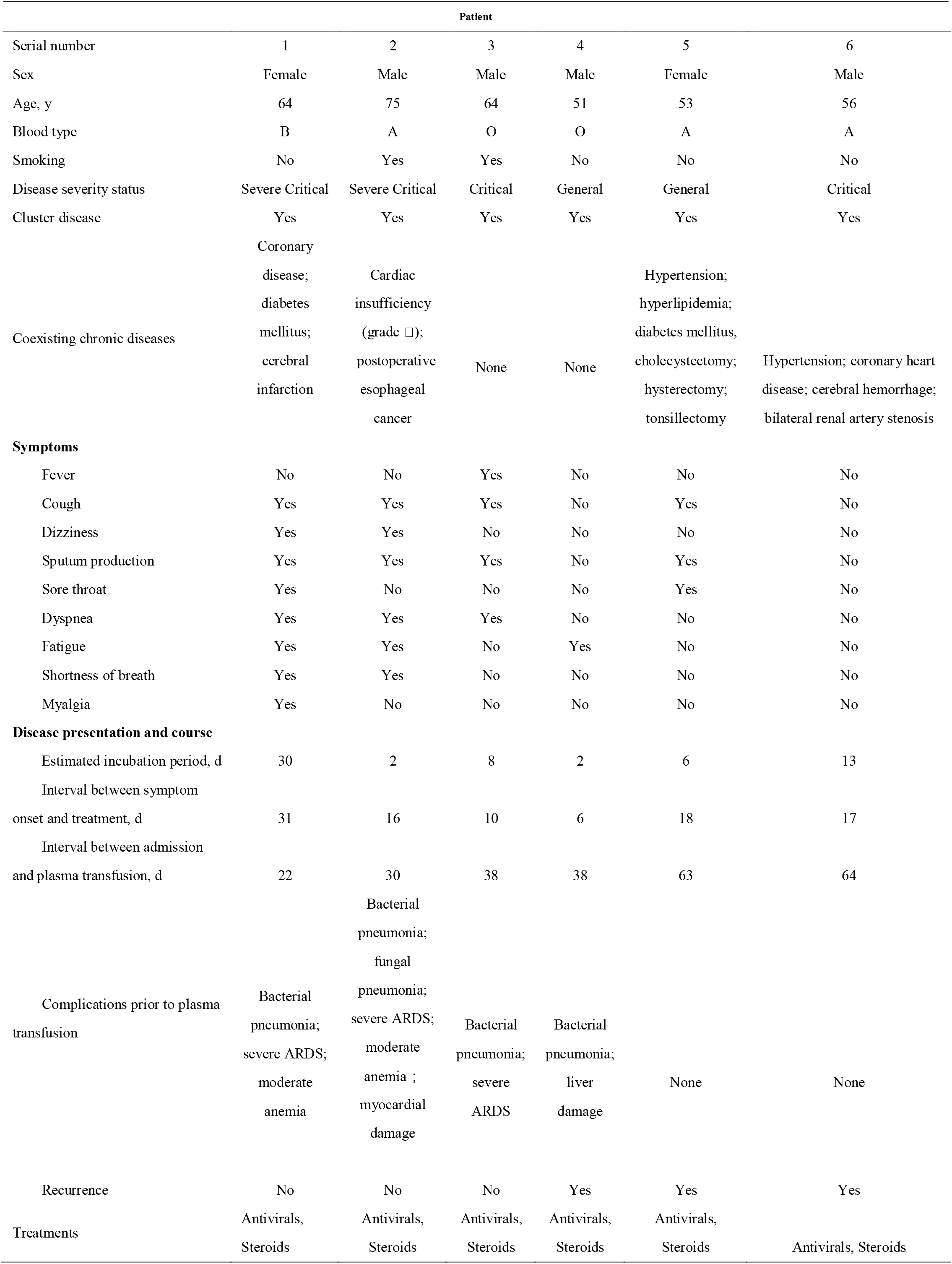
**Clinical characteristics of the 6 participants who received convalescent plasma**

### Donors and convalescent plasma transfusion

Convalescent plasma was collected from six patients who had recovered from COVID-19; their written informed consent was obtained prior to collection. The donors were required to: 1) have been asymptomatic for at least three weeks, 2) be confirmed as negative for SARS-CoV-2 nucleic acid by two consecutive RT-PCR tests, 3) be seronegative for HBV, HCV, and HIV based on virus-specific antibody titers, and 4) have a serum SARS-CoV-2-specific ELISA antibody titer of greater than 1:1000 and a neutralizing antibody titer of higher than 40. Following donation, in accordance with the New Coronavirus Pneumonia Convalescent Plasma Therapy Guidance of China (2^nd^ edition)[14], ABO-compatible convalescent plasma from the donors was transfused into the patients on the same day (200 ml each time), which helps maintain the natural activity of the plasma. Adverse events and serious adverse events related to convalescent plasma transfusion were evaluated by treatment clinicians.

### Role of the funding source

The design, data’s collection, analysis, and interpretation, and draft writing of this report was funded by both Science and Technology Support Plan of Guizhou Province and Science and Technology Plan of Guizhou Province.

### Bias

The clinical data were obtained from case records of Hospital Information System (HIS) in Guizhou Provincial Jiangjunshan Hospital, therefore, there are no bias in this study.

## Results

### Patient characteristics

Of the 146 COVID-19 patients in Guizhou Jiangjunshan Hospital, this study was conducted on six patients who received convalescent plasma therapy. These patients were not allergic to the plasma contents, were negative for hepatitis B virus, hepatitis C virus, and human immunodeficiency virus, and did not have any bacterial infections. Among them, three patients rapidly developed severe disease while their SARS-CoV-2 RT-PCR throat swab results remained persistently positive, and three patients had recurrent cases with worsening symptoms after being empirically treated with antiviral drugs. Notably, Guizhou Jiangjunshan Hospital did not directly receive any COVID-19 patients directly from the community. All patients in this hospital were first treated in other non-designated hospitals for some time before being transferred to Jiangjunshan Hospital. Therefore, these patients were already relatively far along in the course of their illness when admitted to our hospital. Patients 1, 2, and 3 were in the treatment period, and they each had a disease severity status of critical or severe critical. Their conditions were not improved by general antiviral therapy, so they were treated with convalescent plasma transfusion. Patients 4, 5, and 6 were in recurrence; regarding their disease severity, two of them were classified as general, whereas the third was critical. Because of previous treatments, the symptoms, blood cell count, coagulation test results, and biochemical analysis data upon admission for patients 4, 5, and 6 were mostly normal; however, because the laryngeal swab test results were still positive for a long time after disease recurrence, these patients were treated with convalescent plasma therapy. No adverse reactions were observed in any of the six patients during or after the convalescent plasma transfusion.

Two of the patients were smokers, and two had no preexisting medical conditions. All six patients had been infected via contact with other patients diagnosed with COVID-19 and had subsequently taken various antiviral agents and steroids. Most patients experienced cough, dizziness, sputum production, dyspnea, fatigue, and shortness of breath at the onset of disease. Other common clinical manifestations included fever, sore throat, and myalgia. Even though fever is considered to be a particularly important symptom of COVID-19, only one of these six patients experienced a fever. The shortest incubation period was 2 days, and the longest was 30 days. The interval between the initial admission and the convalescent plasma was administered is 22 ∼ 64 days (Table 1).

### Convalescent plasma treatment for the patients with persistent COVID-19

Patient 1 was hospitalized for the first time on February 12 and was directly admitted to Jiangjunshan Hospital. At admission, her oxygen saturation was 85%, she had a PaO_2_/FiO_2_ of 128 (normal range: >300), and she began receiving oxygen therapy through a nasal catheter. Chest imaging on February 24 showed multiple ground glass opacities (GGOs) in both sides of the lungs. Her condition worsened, and she continued to experience respiratory distress and require oxygen supplementation. At this time, although plasma therapy had been planned for patients, we had not yet obtained convalescent plasma that was compatible with the patient’s ABO, so her treatment continued with only antiviral therapy. On March 4, her symptoms and oxygenation were relatively stable. Repeated chest imaging examinations suggested that there were still irregular patch density lesions in both lungs. Therefore, convalescent plasma was initially given on March 4, with a second cycle administered on March 5. After the treatment, the patient’s levels of the inflammatory biomarkers CRP, IL-6, and procalcitonin all decreased. Some resolution of pulmonary lesions was observed at 3 days after the plasma treatment. Meanwhile, her lymphocyte counts gradually returned to normal from their previous low levels, but her monocyte count remained high (10.10–12.40; normal range, 0–7). The interval between administering convalescent plasma therapy and obtaining negative SARS-CoV-2 RT-PCR results for a laryngeal swab was five days (Table 2).

**Table 2.**
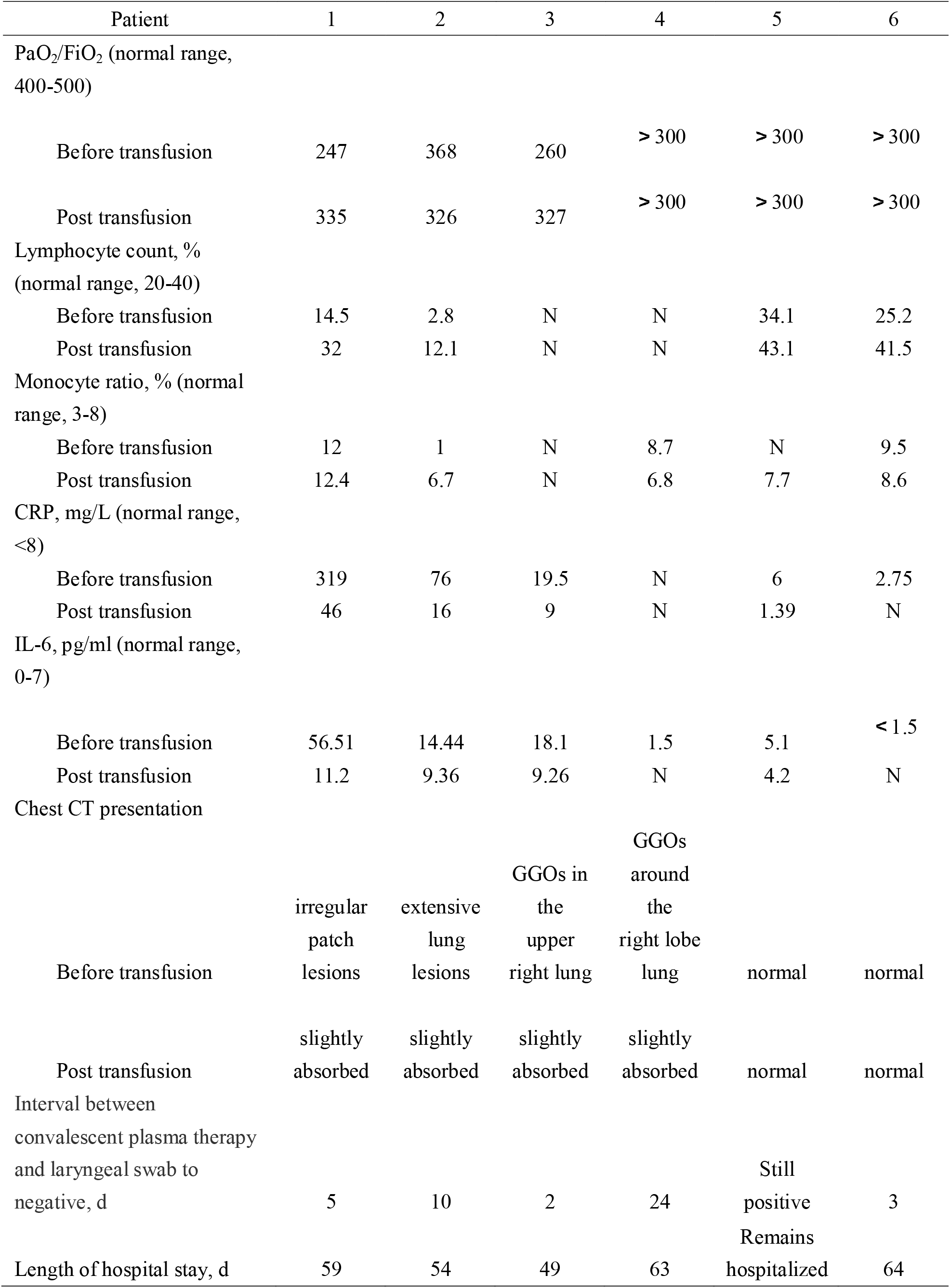
**Comparison of clinical Indexes and laboratory results before and after convalescent plasma transfusion**

Patient 2, a 75-year-old severe critical patient, was first diagnosed and admitted to Jiangjunshan Hospital on February 2. He had elevated levels of CRP (19.5) and IL-6 (18.1), and he was moderately anemic. Chest imaging on March 1 showed no changes in the multiple GGOs in both sides of his lungs and worse lesions in the right lower lobe. Secondary fungal infections were also detected. During this period, SARS-CoV-2 RT-PCR tests on the patient’s throat swabs continued to yield positive results, and the patient continued to have moderate anemia. He received an initial transfusion of convalescent plasma on March 3, followed by a second cycle on March 4. The patient experienced a gradual resolution of his pulmonary lesions after the plasma treatment (Figure 2), and his CRP level, IL-6 level, and neutrophil count gradually decreased from their abnormally high values. Additionally, his lymphocyte counts gradually increased from their abnormally low level, and his monocyte count gradually increased from low to normal. The interval between administering convalescent plasma therapy and obtaining negative SARS-CoV-2 RT-PCR results for a laryngeal swab was ten days (Table 2).

**Figure 1.**
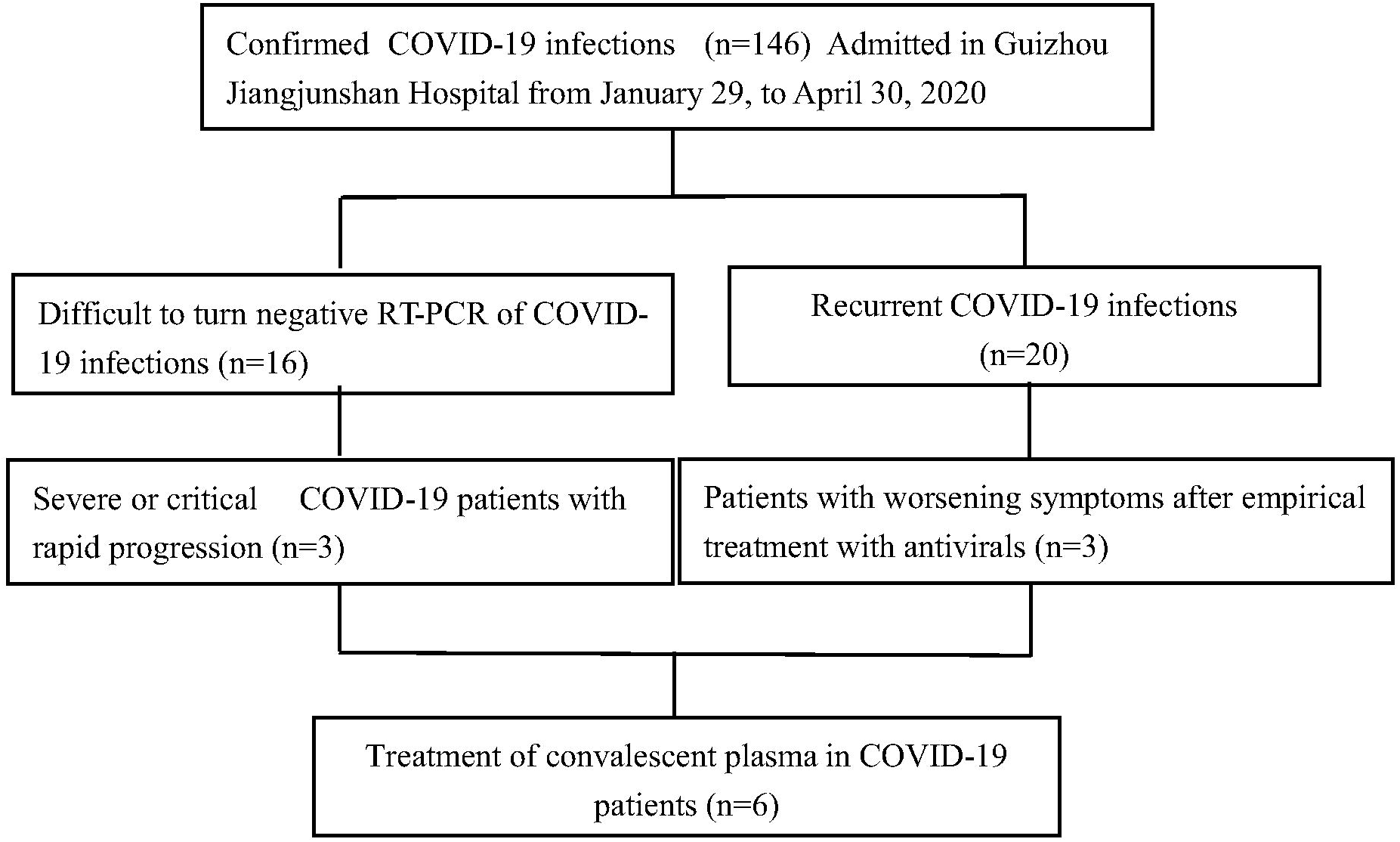
CONSORT Flow Diagram of This Study

**Figure 2.**
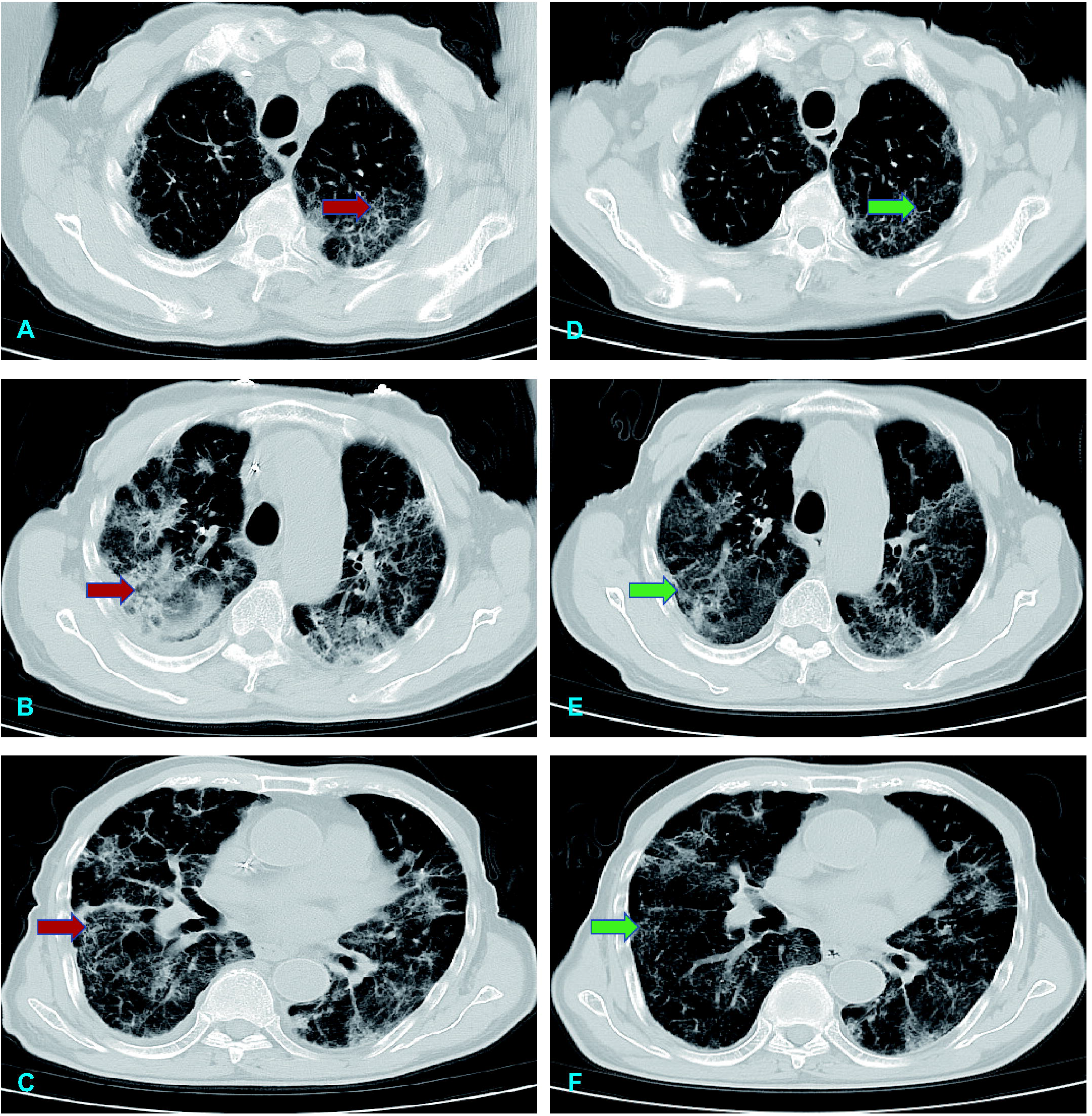
Changes of Chest CT before and after Transfusion of Convalescent Plasma in a 75-year-old Male Patient with Severe COVID-19 Pneumonia. Panel| A, B, and C denote the features before transfusion of covalescent plasma; Panel| D, E, and F denote the features after transfusion of covalescent plasma; Red Arrow denotes inflammatory lesions before transfusion of covalescent plasma; Green Arrow denotes decreased inflammatory lesions after transfusion of covalescent plasma.

Patient 3 was admitted to our hospital on February 4. He continued to have a fever (maximum temperature, 38.5 °C) and PaO_2_/FiO_2_ of < 300 from February 2 up until he received convalescent plasma treatment on March 14. A chest CT on March 5 revealed the sustained presence of GGOs in the upper right lung. Before receiving the convalescent plasma transfusion, the patient’s PaO_2_/FiO_2_ was 260; this value rose to 327 only one day after this intervention. The patient’s body temperature (37.6 °C just before the plasma transfusion) also returned to normal. It took only two days from receiving the convalescent plasma therapy for the patient to get negative SARS-CoV-2 RT-PCR results from a laryngeal swab (Table 2). Additionally, repeated chest CT scans revealed that the plaque density lesions in both sides of the lungs gradually disappeared.

### Convalescent plasma treatment for the patients with COVID-19 recurrence

Patients 4, 5, and 6 had a recurrence of COVID-19. Because they had received previous treatment, they were asymptomatic, and their blood cell count and biochemical analysis data were basically normal at admission. A chest CT of patient 4 revealed GGOs around the right lobe of the lung, whereas the chest CT scans of patients 3 and 5 were normal. The time intervals from the first admission at which patients 4, 5, and 6 were diagnosed with COVID-19 to the first negative laryngeal swab test results were 25, 14, and 18 days, respectively. The time from the first negative laryngeal swab test result to a subsequent positive laryngeal swab test was 13, 47, and 37 days for patients 4, 5, and 6, respectively. The time between the readmission of patients 4 and 5 to the hospital and the adoption of plasma therapy was 3 and 6 days, respectively; these patients received only one round of convalescent plasma transfusion. Patient 6 received convalescent plasma transfusions on the third and 25^th^ days after re-admission. All patients continued receiving antiviral therapy while receiving the convalescent plasma therapy. At 24 days after the initial plasma treatment, patient 4 obtained negative results from a pharyngeal test strip. Patient 6 obtained negative SARS-CoV-2 RT-PCR test results on the third day after receiving convalescent plasma. These two patients were discharged after several consecutive SARS-CoV-2 pharyngeal swab tests produced negative results. As of April 27, 2020, which was 22 days after patient 5 first received plasma therapy, pharyngeal tests on this patient’s still produced positive results, so this patient remained hospitalized.

## Discussion

This descriptive study was conducted on six cases to explore the efficacy of convalescent plasma therapy for treating COVID-19. According to previous experience with convalescent plasma treatment in SARS and severe influenza[15, 16], the production of endogenous IgM and IgG antibodies peaks at two and four weeks, respectively, after infection. In theory, it should be more effective to use convalescent plasma as soon as possible in the early stage of the disease. However, our study differs from previous convalescent plasma studies on SARS and influenza cases in that we used convalescent plasma during the late stage of disease. The long interval from disease onset to plasma treatment in our study was dictated by the type of patients admitted to Guizhou Jianjunshan Hospital; as a tertiary hospital, the admitted patients have been treated elsewhere before being transferred.

Notably, other treatments may affect the relationship between convalescent plasma and antibody levels, such as antiviral drugs and steroids[8]. Fortunately, we found that in all six cases, convalescent plasma was still clinically beneficial. Patient 1 was in critical condition before receiving the convalescent plasma treatment. While waiting for the convalescent plasma for this patient, we continued to treat her with antiviral drugs and steroids, which clearly stabilized the disease condition, so these treatments likely also contributed to her recovery. There has been much debate about the use of steroids, and it has not been used as a routine treatment from the beginning. Evidence from SARS shows that corticosteroid treatment cannot reduce mortality and it can delay virus clearance[17]. However, systemic corticosteroids may help treat COVID-19 by reducing the excessive lung damage caused by inflammation[18]. The conditions of the six patients in our study had all rapidly deteriorated to ARDS, so they were given intravenous methylprednisolone before convalescent plasma therapy. After patient 1 was given steroid therapy, but before she was given the convalescent plasma, her PaO_2_/FiO_2_ underwent a rapid improvement (from 128 to 247), although it did not return to a normal level. After the convalescent plasma treatment, her PaO_2_/FiO_2_ further increased to 335. After they were treated with convalescent plasma, the PaO_2_/FiO_2_ and lymphocyte count of patients 1, 2, and 3 were close to normal. Additionally, the levels of inflammation markers CRP and IL-6 for these patients decreased significantly, and chest imaging examinations revealed that their lung lesions gradually subsided. The autopsy reports for COVID-19 fatalities are very similar to those for SARS-CoV and MERS-CoV fatalities; they describe diffuse alveolar injury, exudate, and inflammation[19]. ARDS in these patients is partially caused by the cytokine storm and the host immune response[20]. Therefore, the anti-SARS-CoV-2 IgM and IgG in the convalescent plasma treatment not only can directly neutralize the virus, but these anti-inflammatory components can also accelerate the clearance of infected cells to prevent cytokine storms. It is not surprising that for the patients with recurrence of COVID-19, the minimum time for viral clearance (as assessed by two consecutive negative throat swab tests for SARS-CoV-2 nucleic acid) after the treatment with convalescent plasma is 2 days, and the maximum time is 24 days. However, patient 5 has had consistently positive laryngeal swab test results since receiving convalescent plasma on April 5, 2020 up through the time of writing, May 20, 2020. One possible reason is that the patient’s basic condition and immune system have been poor because this patient suffers from high blood pressure, hyperlipidemia, and type 2 diabetes, and the patient has undergone various operations (e.g., cholecystectomy, hysterectomy, and tonsillectomy). Before infusion of convalescent plasma during the recovery period, his IgM and IgG levels were abnormally low (0.8 g/L and 5.56 g/L, respectively), and his CD4^+^/CD8^+^ ratio was 4.34. Another possibility is that the anti-SARS-CoV-2 IgM and IgG in the convalescent plasma treatment were insufficient to neutralize the virus in this patient’s body. Notably, this patient received only one round of convalescent plasma therapy, after which he was treated with antiviral drugs and steroids. We speculate that if another round of convalescent plasma transfusion is given to this patient, the effect of this intervention could be enhanced, speeding up the time until negative results are obtained for laryngeal swab tests of SARS-CoV-2 nucleic acid. Furthermore, a recent report found that antibodies in convalescent plasma administered during the recovery period can help maintain a higher antibody titer and increase the host immune response until the virus infection has cleared[5].

This study was limited by its small sample size. However, our study population was typical of patients in Guizhou Province, and it included both patients who were treated with plasma therapy during their first SARS-CoV-2 infection and COVID-19 patients who relapsed. Importantly, the treatment of COVID-19 patients with convalescent plasma did not cause any serious adverse effects.

In sum up, six patients with COVID-19 were treated with antiviral drugs and systemic corticosteroids combined with appropriate rounds of convalescent plasma therapy, after which their clinical conditions were effectively improved. Thus, transfusion with convalescent plasma is another viable option for the treatment of COVID-19, especially when used in combination with antiviral drugs and systemic corticosteroids. As more and more COVID-19 patients have recovered from the infection, convalescent plasma therapy can be encouraged, and active voluntary donation of convalescent plasma will be greatly appreciated.

## Data Availability

We stated that all the data and materials were true and available in the study.

http://www.chictr.org.cn/edit.aspx?pid=53859&htm=4

## Acknowledgements

We thank Ping Lu, Min Yao, Yang Hong, and Jinhua Wu for providing selfless help with the process of database construction for COVID-19 in this study. We also thank Katie Oakley, PhD, from Liwen Bianji, Edanz Editing China (www.liwenbianji.cn/ac), for editing the English text of a draft of this manuscript.

## Authorship contributions

Wei Zhang had full access to all data in the present study and accepts responsibility for data management and accuracy of the data analyses. Study concept and design: Wei Zhang and Weidong Wu. Acquisition and interpretation of data: Can Jin, Juan Gu, and Youshu Yuan. Drafting of the manuscript: Can Jin and Wei Zhang. Critical revision of the manuscript for important intellectual content: Weidong Wu and Wei Zhang. Administrative, technical, or material support: Qinying Long, Qi Zhang, and Hourong Zhou. Study supervision: Wei Zhang and Weidong Wu. All authors agree to submission of the final version of this manuscript. Wei Zhang is the study guarantor.

## Disclosure of Conflicts of Interest

All authors have no conflicts of interest to disclose.

## Funding

This study was supported by Science and Technology Support Plan of Guizhou Province in 2019 (Qian Ke He Support [2019] 2834) and Science and Technology Plan of Guizhou Province in 2020 (Qian Ke He Fundamental [2020] 1Z061).

## Figure legends

Figure 1. CONSORT flow diagram of this study

Figure 2. Chest CT scans of a 75-year-old male patient with severe COVID-19 pneumonia before and after transfusion with convalescent plasma

## Notes

### Competing Interest Statement

The authors have declared no competing interest.

### Clinical Trial

ChiCTR2000033056

### Clinical Protocols

http://www.chictr.org.cn/edit.aspx?pid=53859&htm=4

### Author Declarations

This study was approved by the Biomedical Ethics Committee of Affiliated Hospital of Zunyi Medical University.

